# Leveraging Real-World Evidence to Define Severe RSV Lower Respiratory Tract Disease in Adults

**DOI:** 10.1101/2024.03.20.24304618

**Authors:** Catherine A. Panozzo, Edward E. Walsh, Zhen Yang, Eleanor Wilson, Jaya Goswami, Sonia K. Stoszek, Adrianna Loback, Tony Ng, Beverly M. Francis, Alana K. Simorellis, Wenmei Huang, Linwei Li, Rebecca Vislay-Wade, Zhe Zheng, Evan J. Anderson, Allison August, Grace Chen, Ann R. Falsey

## Abstract

This study analyzed previously published data in hospital and community cohorts of adults with respiratory syncytial virus (RSV)–associated symptoms. Shortness of breath (dyspnea) alone, and in combination with certain other lower respiratory tract disease signs/symptoms, was a leading symptomatic indicator for severe RSV outcomes.

## INTRODUCTION

Respiratory syncytial virus (RSV) affects people of all ages; older adults and individuals with certain underlying conditions are at higher risk of developing complications from RSV disease [1-3]. RSV typically causes mild upper respiratory tract symptoms [3, 4], but can progress to severe lower respiratory tract disease (LRTD) in older and at-risk adults, potentially leading to pneumonia, hospitalization, and death [3, 4]. In 2019, RSV caused an estimated 5.2 million cases, 466,000 hospitalizations, and 33,000 in-hospital deaths among high-income countries in adults aged ≥60 years [5].

Laboratory testing is necessary to confirm an RSV diagnosis, as RSV is not easily clinically distinguishable from other respiratory pathogens [2]. In addition, there are no specific agreed-upon signs and symptoms to distinguish RSV-acute respiratory disease (ARD) from RSV-LRTD, or severe RSV-LRTD. Consequently, no standard case definitions exist for these outcomes in adults [2, 5, 6]. This, in addition to an overall lower sensitivity of diagnostic tests and infrequent testing in adults [5, 7], may contribute to an underestimation of RSV-associated disease and present challenges when comparing results across vaccine and therapeutic studies.

This study investigated definitions of severe RSV-LRTD that could be leveraged for the analyses of adult RSV vaccine efficacy trials.

## METHODS

### Population

The study retrospectively analyzed data from a cohort study of adults aged ≥18 years (median age, 73 years; Interquartile range, 18 years) who were evaluated for signs/symptoms of ARD and tested for RSV by reverse transcription-polymerase chain reaction in Rochester, New York [8]. The population analyzed included two cohorts: (1) individuals hospitalized with ≥1 ARD sign/symptom or an acute cardiopulmonary illness (hospitalized cohort), and (2) individuals enrolled and followed prospectively for ARD in the community (including those who did and did not seek medical care for ARD; community cohort). Additional population selection criteria are shown in the **Supplemental Methods**.

### Probability of Individual or Paired ARD Signs/Symptoms

This analysis assessed the probability with which symptoms or combinations of symptoms were associated with positive versus negative RSV diagnoses in hospitalized versus community cohorts (e.g., proxies for severe vs less severe RSV disease), and in medically attended versus non–medically attended individuals (approximating moderate vs mild RSV disease). A z-test was used to infer statistical differences between the frequencies (additional details are in **Supplemental Methods**).

### Assessment of Severe RSV-LRTD Case Definitions

Informed by the frequencies of signs/symptoms observed across a spectrum of RSV disease, we evaluated the performance of proposed severe RSV-LRTD case definitions in predicting hospitalized versus community cases, as well as medically attended versus non–medically attended community cases, by applying them to the dataset.

### Sensitivity and Specificity Analysis

Sensitivity and specificity of case definitions were calculated to accurately identify severe RSV-LRTD cases (described in **Supplemental Methods**). A machine learning approach was also used to predict severe RSV-LRTD among RSV-positive patients. Logistic regression with ridge regularization and Explainable Boosting Machine [9] models were trained using various input variables (**Supplemental Table 1**).

## RESULTS

### Participant Characteristics

The data retrospectively analyzed from the original study by Falsey et al [8] included 999 symptomatic individuals, with 163 confirmed cases of RSV (RSV-positive) and 836 without RSV (RSV-negative; **Supplemental Figure 1**), as confirmed by reverse transcription-polymerase chain reaction. Among the 836 RSV-negative participants, 156 (18.6%) had evidence of another viral respiratory pathogen (48.1% had influenza A [n = 75] and 23.7% had human metapneumovirus [n = 37]). Both RSV-positive and RSV-negative groups were similar with respect to age and the proportion of participants reporting underlying chronic obstructive pulmonary disease, congestive heart failure, diabetes mellitus, renal failure, average baseline respiratory rate, and oxygen saturation (**Supplemental Table 2**). Among the 22 RSV-positive participants in the community cohort who sought medical care, 19 received care in the outpatient setting, 2 received care in both the emergency department and hospital setting, and 1 received care in both the outpatient and hospital setting. As these individuals were not initially hospitalized, they were included in the community cohort.

### Comparison of Signs/Symptoms

#### RSV-Positive Versus RSV-Negative Cases

Overall, cough (90%), nasal congestion (79%), sputum production (72%), and abnormal lung sounds by auscultation (71%) were the most common signs/symptoms among RSV-positive participants (n = 163). Compared with RSV-negative participants (n = 836), RSV-positive participants more frequently reported wheezing or sputum production or had rhonchi on examination (**Supplemental Figure 2A**).

#### RSV-Positive Cases: Hospitalized Cohort Versus Community Cohort

Shortness of breath (dyspnea; 98%), abnormal lung sounds by auscultation (98%), cough (94%), and tachypnea (91%) were the most common signs/symptoms among hospitalized RSV-positive patients (n = 85). A greater percentage of hospitalized RSV-positive patients reported dyspnea, hypoxemia, tachypnea, rales, rhonchi, fever, or wheezing compared with those in the community cohort (n = 78; **Supplemental Figure 2B**).

#### Community Cohort RSV-Positive Cases: Medically Attended Versus Non–Medically Attended

For medically attended RSV-positive participants in the community cohort (i.e., ≥1 visit to the outpatient, emergency department, or inpatient setting; n = 22), cough (100%), sputum production (86%), and nasal congestion (82%) were the most reported ARD symptoms. A greater percentage of medically attended RSV-positive participants in the community cohort had sputum production, hypoxemia, or cough than non–medically attended RSV-positive participants in the community cohort. Dyspnea was observed in 41% of medically attended RSV-positive participants compared to 23% of non– medically attended RSV-positive participants (n = 56; **Supplemental Figure 2C**).

#### Sensitivity and Specificity Analyses of Severe RSV-LRTD Case Definitions

The highest sensitivity (100%) was observed with any combination of 2 of the following signs/symptoms: dyspnea, abnormal lung sounds by auscultation, fever and/or cough, hypoxemia, or tachypnea (**Figure 1; Supplemental Table 3**); however, this was at the expense of specificity (46%). The definitions demonstrating the best balance between sensitivity (98%) and specificity (72%-76%) were those that included dyspnea alone, or either of the following variations: 1) dyspnea AND abnormal lung sounds by auscultation OR hypoxemia OR tachypnea; 2) dyspnea AND abnormal lung sounds by auscultation OR cough and/or fever; 3) dyspnea AND abnormal lung sounds by auscultation OR cough and/or fever OR hypoxemia OR tachypnea; 4) dyspnea AND abnormal lung sounds by auscultation, cough and/or fever, sputum production, chest pain, tachypnea, hypoxemia. Cough and sputum production were not specific for hospitalization compared with other definitions that may or may not have included cough. Probability of paired ARD signs/symptoms is discussed in the Supplemental Results.

**Figure 1.**
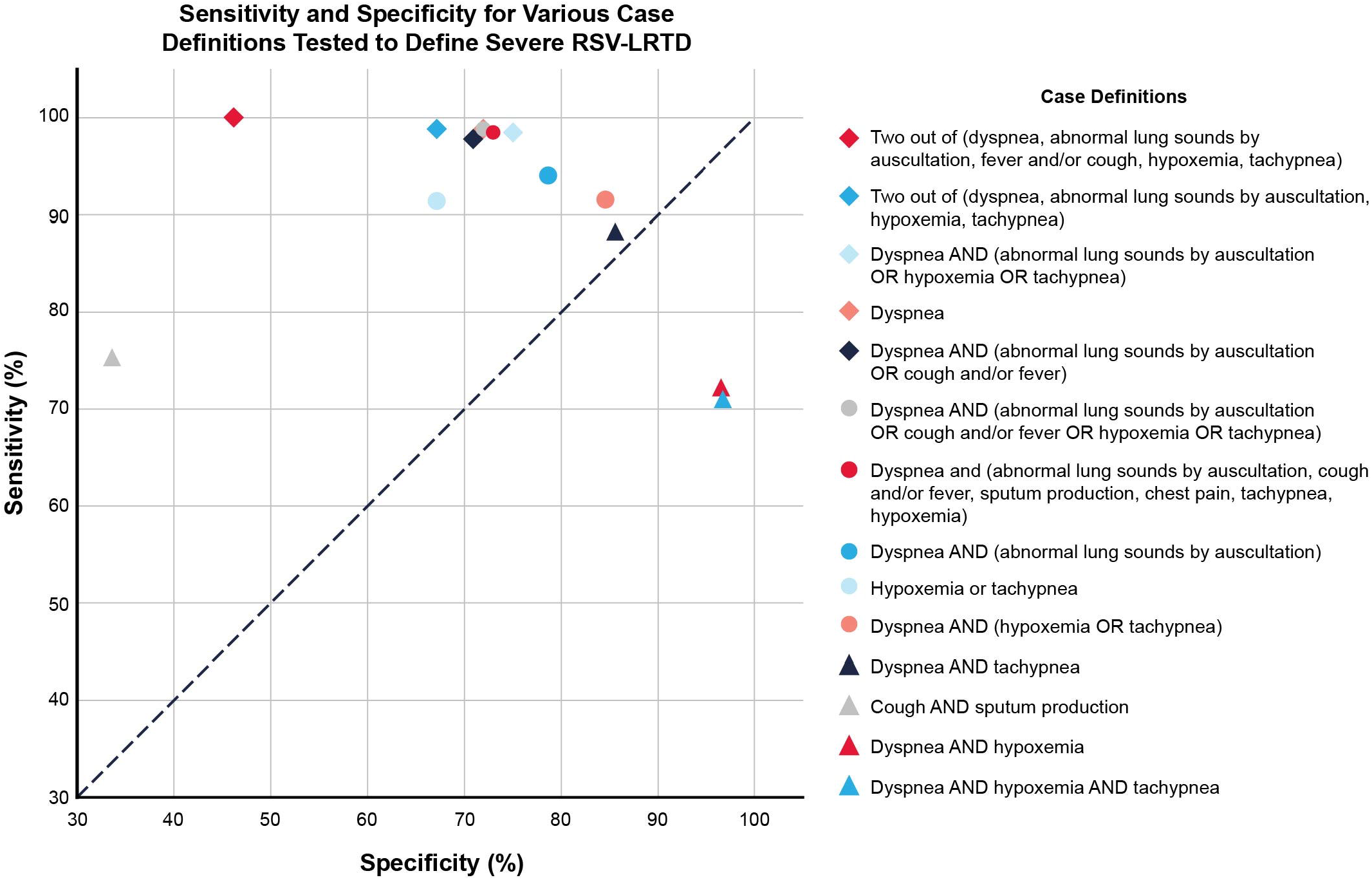
Assessment of sensitivity and specificity across different case definitions for severe RSV-LRTD classification. Abnormal lung sounds by auscultation refers to wheeze/rales/rhonchi. Abbreviations: LRTD, lower respiratory tract disease; RSV, respiratory syncytial virus.

#### Sensitivity and Specificity Analyses of Severe RSV-LRTD Using a Machine Learning Approach

A machine learning approach to predict severe RSV-LRTD (e.g., hospitalization from RSV) had a sensitivity of 1.0 and specificity of 0.80 for logistic regression, and a sensitivity of 0.94 and specificity of 0.86 for Explainable Boosting Machine (**Supplemental Table 4**; additional information available in **Supplemental Results**). Consistent with results from the non–machine learning approach, the variables with the largest weights (i.e., ranked as most important in predicting hospitalization) were dyspnea (**Supplemental Figure 3**), followed by hypoxemia, tachypnea, and fever. Other variables that were found to impact risk of hospitalization included age, pulse, rales, and combination of dyspnea and body temperature.

## DISCUSSION

This is the first study to our knowledge to reanalyze data from a prospective observational cohort to develop case definitions for severe RSV-LRTD for adults. Dyspnea as a symptom alone had a notably high sensitivity and specificity for predicting severe outcomes (i.e., hospitalization). Across the spectrum of increasingly severe RSV disease, the frequency of dyspnea increased from 23% in the RSV non–medically attended community cohort to 41% in the RSV medically attended community cohort to 98% in the hospitalized cohort. Given the high sensitivity of dyspnea alone for predicting RSV-LRTD–associated hospitalizations in our analysis, this symptom should be considered as a leading symptomatic indicator of LRTD and severe RSV disease in future studies. As other studies have suggested, the analyses showed that no individual or combination of signs/symptoms were highly specific in distinguishing between RSV-positive and RSV-negative individuals, although there were general trends [2]. Hence, laboratory testing combined with a comprehensive list of LRTD signs/symptoms are critical features of any RSV-LRTD case definition.

This study had several limitations, including a relatively small sample size in certain groups, limited geographic scope, and a population that included a high proportion of individuals with COPD and self-reported smokers, thus limiting its generalizability. Study strengths included prospective and systematic collection of signs and symptoms across both hospital and community cohorts, and the collection of data over 4 RSV seasons.

This study demonstrates how data from a well-characterized observational cohort can inform case definitions in a clinical trial. Additionally, this study provides information regarding which signs and symptoms of RSV disease are related to healthcare-seeking behavior and hospitalizations in adult cohorts, where markers for severe RSV disease are not standardized. As the study population included adults of all ages, these data can be considered in developing case definitions for severe RSV-LRTD across the adult lifespan, including older adults and younger high-risk populations.

## Data Availability

The authors declare that the data supporting the findings of this study are available within this article.

## Acknowledgements

Medical writing and editorial assistance was provided by Meenu Minhas, PhD, of MEDiSTRAVA in accordance with Good Publication Practice (GPP3) guidelines, funded by Moderna, Inc., and under the direction of the authors.

## Author contributions

CP designed the concept. CP, EW, ZY, EW, JG, SS, AL, TN, BMF, AS, LL, EJA, and AF analyzed or interpreted the data. All authors provided writing, review, or intellectual contributions and approved the final draft

