## Supplementary Material for "Leveraging Real-World Evidence to Define Severe RSV Lower Respiratory Tract Disease in Adults"

### Supplemental Materials

### Supplemental Methods

#### *Population*

The current study reanalyzed data from an existing cohort study that prospectively evaluated participants aged ≥18 years (median age: 73 years; Interquartile range, 18 years) for signs/symptoms (nasal congestion, sore throat, hoarseness, new or worsening cough, sputum production, and shortness of breath [dyspnea] with or without fever) of acute respiratory disease (ARD), and performed testing of respiratory syncytial virus (RSV) by reverse transcription-polymerase chain reaction (RT-PCR) over 4 seasons (1999-2003) in Rochester, New York (Falsey et al [1]). Seropositive-only cases were not included. The population analyzed consisted of 2 cohorts based on how they were enrolled: persons admitted to the hospital with ≥1 ARD sign/symptom or an acute cardiopulmonary illness (hospitalized cohort), or persons enrolled and followed prospectively for ARD in the community (includes persons who did and did not seek medical care for ARD; community cohort; **Supplemental** **Figure 1**). Participants from these cohorts were included in the current study if they had a documented symptomatic ARD and a corresponding positive or negative RSV RT-PCR test result. The probability of ARD signs/symptoms was described alone and in pairs for RSV-positive and RSV-negative cases and were stratified by hospitalized and community cohorts.

The analyses in this report utilized adaptations of the above definitions based on data availability from the Falsey et al [1] study. The modified definition of RSV-LRTD used in this study includes the presence of new or worsening of ≥2 or ≥3 of the following signs/symptoms for ≥24 hours: dyspnea, cough and/or fever (≥37.8°C [100.0°F]), abnormal lung sounds by auscultation (wheezing or rales or rhonchi [wheezing is defined as wheezing from physical examination findings or self-reported wheezing]), sputum production, tachypnea (respiratory rate ≥20 breaths per minute), hypoxemia (oxygen saturation ≤93%), and pleuritic chest pain.

#### *Probability of Paired ARD Signs/Symptoms*

The first set of analyses included a z test assessment to help characterize signs/symptoms, or combinations of signs/symptoms, more commonly associated with positive versus negative RSV diagnoses, hospitalized versus community cohorts (i.e., proxies for more vs less severe RSV disease) and medically attended vs non–medically attended patients (i.e., also to approximate more vs less severe RSV disease). Medically attended analyses were limited to RSV-positive participants in the community cohort, and a medically attended event was defined as having ≥1 visit to an outpatient, emergency department, or inpatient setting.

*Sensitivity and Specificity Analysis*

The performance of selected case definitions was evaluated based on sensitivity and specificity of case definitions to accurately identify RSV-LRTD. In the absence of a standardized case definition for severe RSV-LRTD, signs/symptoms experienced by hospitalized RSV-positive patients were considered as a proxy for signs/symptoms of severe RSV-LRTD, since hospitalization indicates a more severe manifestation of the disease. Similarly, signs/symptoms experienced by medically attended RSV-positive patients in the community cohort were considered a proxy for moderate to severe RSV-LRTD, since seeking any kind of medical care (e.g., in the outpatient setting) may indicate more severe disease. Sensitivity and specificity were calculated using the following equations: sensitivity = [true positive/(true positive + false negative)]*100; specificity = [true negative/( true negative + false positive)]*100 [2].

#### *Machine Learning Approach to Identify Severe RSV-LRTD*

A machine learning approach was employed to predict severe RSV-LRTD (e.g., hospitalization cohort) among all patients testing positive for RSV across the hospital (n = 85) and community cohorts (n = 78). The RSV-positive cohort (n = 163) was randomly split into an 80% training set (n = 130) and a 20% test set (n = 33). Logistic regression with ridge regularization and Explainable Boosting Machine [3, 4] were trained using 24 input variables, including the variables used to define ARD signs/symptoms described previously, and expanded to include variables related to demographics, comorbidities, and vital signs (**Supplemental Table 1**). As machine learning approaches are data driven, the expanded variable list provides additional insight into other important determinants of severe RSV-LRTD disease. To maintain maximum information, continuous variables previously categorized as binary were entered into the model as continuous. Due to sample size limitations, pairwise interactions were not considered. The machine learning analyses reported sensitivity, specificity, and variable importance as a comparison to results of the non–machine learning approach.

### Supplemental Results

#### **Probability of Paired ARD Signs/Symptoms**

##### *RSV-Positive Versus RSV-Negative Cases*

In the combined hospitalized and community cohorts, the most common pairs of ARD symptoms for RSV-positive patients (n = 163) were cough + sputum production (72%), cough + nasal congestion (71%), cough + abnormal lung sounds by auscultation (67%), cough + dyspnea (61%), cough + tachypnea (58%), and cough + wheezing (58%) (**Supplemental Figure 4A**). Similar combinations of symptom pairs were observed for RSV-negative patients (n = 836; **Supplemental Figure 4B**). Combinations of tachypnea + wheezing (17%), cough + wheezing (16%), abnormal lung sounds by auscultation + wheezing (16%), cough + sputum production (16%), and dyspnea + rhonchi (16%) best distinguished between RSV-positive and RSV-negative cases; however, differences were small (**Supplemental** **Figure 5A**).

##### *RSV-Positive Cases: Hospitalized Cohort Versus Community Cohort*

The most common pairs of ARD symptoms for RSV-positive hospitalized patients were dyspnea + abnormal lung sounds by auscultation (95%), cough + abnormal lung sounds by auscultation (93%), dyspnea + cough (92%), dyspnea + tachypnea (89%), and tachypnea + abnormal lung sounds by auscultation (88%) (**Supplemental Figure 4C**). The RSV-positive community cohort most commonly experienced the following symptom pairs: cough + nasal congestion (78%) and nasal congestion + rhinorrhea (74%) (**Supplemental Figure 4D**). Combinations of dyspnea + tachypnea (74%), dyspnea + abnormal lung sounds by auscultation (73%), dyspnea + hypoxemia (68%), and tachypnea + abnormal lung sounds by auscultation (67%) best distinguished between RSV-positive hospitalized and RSV-positive community cases (**Supplemental** **Figure 5B**).

##### *Community Cohort RSV-Positive Cases: Medically Attended Versus Non–Medically Attended*

The most common pairs of ARD symptoms for RSV-positive patients seeking medical care in the community cohort were cough + sputum production (86%) or cough + nasal congestion (82%) (**Supplemental Figure 4E**). For RSV-positive patients not seeking medical care in the community cohort, the most common paired signs/symptoms were cough + nasal congestion (77%) and nasal congestion + rhinorrhea (77%; **Supplemental Figure 4F**). The combinations of hoarseness + sputum production (30%), tachypnea + hypoxemia (27%), and cough + sputum production (27%) production best distinguished between medically attended or non–medically attended RSV-positive patients in the community cohort (**Supplemental** **Figure 5C**).

**Machine Learning Approach**

Eleven of the top 12 most important variables in predicting severe RSV-LRTD were signs and symptoms. Dyspnea followed by oxygen saturation, respiratory rate, and temperature were most predictive of severe RSV-LRTD. Age was the fifth most important predictive variable and may have been bolstered by the fact that this study included both younger and older adults.

**Supplemental Tables**

#### **Supplemental Table 1. Input Variables for Machine Learning Models**

| **Variable Type** | **Variable Name** |
| --- | --- |
| Continuous | Age, pulse, temperature, oxygen saturation, respiratory rate |
| Categorical | Sex, chronic obstructive pulmonary disease, congestive heart failure, diabetes, renal failure, smoking status, chest pain, constitutional, cough, feverish, hoarseness, nasal congestion, rales, rhinorrhea, rhonchi, dyspnea, sore throat, sputum production, wheezing |

#### **Supplemental Table 2. Description of Analytic Cohorts**

|  | **Overall RSV-Positive and -Negative**  **(n = 999)** | **Hospitalized and Community Cohorts, RSV-Positive**  **(n = 163)** | **Hospitalized and Community Cohorts, RSV-Negative**  **(n = 836)** | **Hospitalized Cohort, RSV-Positive**  **(n = 85)** | **Community Cohort, RSV-Positive**  **(n = 78)** | **MA RSV-Positive, Community Cohort**  **(n = 22)** | **Non–MA RSV-Positive, Community Cohort**  **(n = 56)** |
| --- | --- | --- | --- | --- | --- | --- | --- |
| **Mean age (SD)** | 69.6 (15.2) | 70.0 (16.1) | 69.5 (15.0) | 74.7 (12.4) | 65.0 (18.2) | 69.7 (14.8) | 63.1 (19.2) |
| **Median age (IQR)** | 73.0 (18.0) | 73.0 (19.5) | 73.0 (18) | 78.0 (20.0) | 69.0 (33.5) | 69 (10) | 66.5 (39.5) |
| **Age range** | 19-90 | 19-90 | 19-90 | 45-90 | 19-90 | 36-90 | 19-85 |
| **Female (%)** | 542 (54.3) | 100 (61.3) | 442 (52.9) | 57 (67.1) | 43 (55.1) | 10 (45.5) | 33 (58.9) |
| **COPD (%)** | 400 (40.0) | 55 (33.7) | 345 (41.3) | 41 (48.2) | 14 (17.9) | 7 (31.8) | 7 (12.5) |
| **CHF (%)** | 188 (18.8) | 31 (19.0) | 157 (18.8) | 25 (29.4) | 6 (7.7) | 4 (18.2) | 2 (3.6) |
| **Diabetes mellitus (%)** | 189 (18.9) | 34 (20.9) | 155 (18.5) | 21 (24.7) | 13 (16.7) | 5 (22.7) | 8 (14.3) |
| **Renal failure (%)** | 15 (1.5) | 2 (1.2) | 13 (1.6) | 2 (2.4) | 0 | 0 | 0 |
| **Smoking (%)** | 697 (69.8) | 100 (61.3) | 597 (71.4) | 56 (65.9) | 44 (56.4) | 16 (72.7) | 28 (50.0) |
| **Respiratory rate, mean (SD)** | 21.4 (7.7) | 22.5 (8.0) | 21.2 (7.7) | 27.3 (6.6) | 17.2 (5.8) | 20.1 (7.1) | 16.0 (4.8) |
| **Percentage oxygen saturation, mean (SD)** | 93.0% (6.9) | 92.1% (8.0) | 93.2% (6.7) | 88.4% (8.7) | 96.2% (4.3) | 94.2% (7.3) | 97.0% (1.7) |

Abbreviations: CHF, chronic heart failure; COPD, chronic obstructive pulmonary disease; IQR, interquartile range; MA, medically attended; RSV, respiratory syncytial virus; SD, standard deviation.

#### **Supplemental Table 3. Sensitivity and Specificity for Various Case Definitions Tested to Define Severe RSV-LRTD**^a^

| **Case Definition for Severe RSV-LRTD^a^** | **Sensitivity, % (n/N)^b^** | **Specificity, % (n/N)^b^** |
| --- | --- | --- |
| 2 out of (dyspnea, abnormal lung sounds by auscultation,^c^ fever and/or cough, hypoxemia, tachypnea) | 100  (85/85) | 46  (36/78) |
| 2 out of (dyspnea, abnormal lung sounds by auscultation,^c^ hypoxemia, tachypnea) | 99  (84/85) | 67  (52/78) |
| Dyspnea AND (abnormal lung sounds by auscultation^c^ OR hypoxemia OR tachypnea) | 98  (83/85) | 76  (61/78) |
| Dyspnea | 98  (83/85) | 72  (56/78) |
| Dyspnea AND (abnormal lung sounds by auscultation^c^ OR cough and/or fever) | 98  (83/85) | 72  (56/78) |
| Dyspnea AND (abnormal lung sounds by auscultation^c^ OR cough and/or fever OR hypoxemia OR tachypnea) | 98  (83/85) | 72  (56/78) |
| Dyspnea AND (abnormal lung sounds by auscultation,^c^ cough and/or fever, sputum production, chest pain, tachypnea, hypoxemia) | 98  (83/85) | 72  (56/78) |
| Dyspnea AND (abnormal lung sounds by auscultation^c^) | 95  (81/85) | 78  (61/78) |
| Hypoxemia or tachypnea | 92  (78/85) | 67  (52/78) |
| Dyspnea AND (hypoxemia OR tachypnea) | 91  (77/85) | 85  (66/78) |
| Dyspnea AND tachypnea | 89  (76/85) | 85  (66/78) |
| Cough AND sputum production | 76  (65/85) | 33  (26/78) |
| Dyspnea AND hypoxemia | 72  (61/85) | 96  (75/78) |
| Dyspnea AND hypoxemia AND tachypnea | 71  (60/85) | 96  (75/78) |

Abbreviations: LRTD, lower respiratory tract disease; RSV, respiratory syncytial virus.

^a^Sensitivity and specificity are used to predict RSV hospitalization.

^b^RSV-positive hospitalized, n = 85; RSV-positive community cohort, n = 78.

^c^Abnormal lung sounds by auscultation refers to wheeze/rales/rhonchi.

#### **Supplemental Table 4. Sensitivity and Specificity for Machine Learning Models to Define Severe RSV-LRTD**

| **Model** | **Sensitivity,^a^ %** | **Specificity,^a^ %** |
| --- | --- | --- |
| Logistic regression | 100 | 80 |
| Explainable Boosting Machine^b^ | 94 | 86 |

Abbreviations: LRTD, lower respiratory tract machine; RSV, respiratory syncytial virus.

^a^The sensitivity and specificity in the table was achieved with a default prediction threshold of 0.5, the Explainable Boosting Machine reaches a sensitivity of 100% and specificity of 80%, with a prediction threshold of 0.05.

^b^An Explainable Boosting Machine is a machine learning approach that uses a tree-based, cyclic gradient boosting Generalized Additive Model with automatic interaction detection.

### Supplemental Figures

#### **Supplemental Figure 1.** Participants from Falsey et al, 2005 [1] were reanalyzed based on RSV RT-PCR results, whether they were identified in hospital or community settings, and within the community setting, whether participants went on to seek medical attendance in the outpatient clinic, emergency department, or hospital.

Abbreviations: ARD, acute respiratory disease; RSV, respiratory syncytial virus; RT-PCR, reverse transcription polymerase chain reaction.

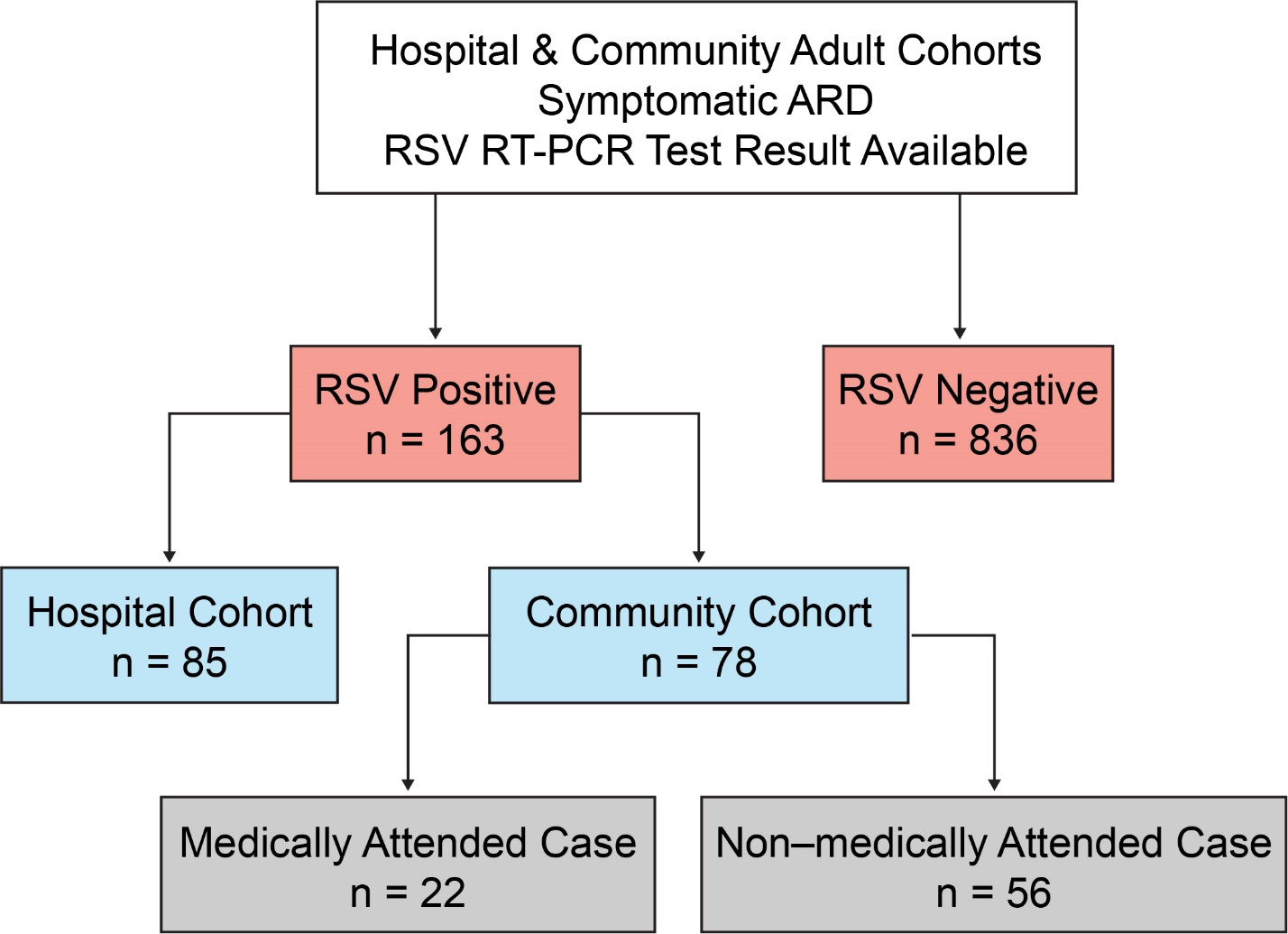

#### **Supplemental Figure 2.** Probability of symptoms for participants. (A) RSV-positive (n = 163) or RSV-negative (n = 836) by RT-PCR; (B) RSV-positive by RT-PCR in the hospitalized (n = 85) or community (n = 78) cohorts; (C) RSV-positive by RT-PCR and medically attended (n = 22) or non–medically attended (n = 56) in the community cohort.

Within each panel, the top color legend represents the probability and difference in frequency of signs/symptoms as percentages. The color scale of probability ranges from 0% to 100%, with darker colors indicating higher percentages. The color scale of difference in probability ranges from −100% to 100%. Positive differences are shown in red and negative differences are shown in blue. The darker the red color, the higher the positive difference, while the darker the blue color, the lower the negative difference.

Abbreviations: RSV, respiratory syncytial virus; RT-PCR; reverse transcription polymerase chain reaction.

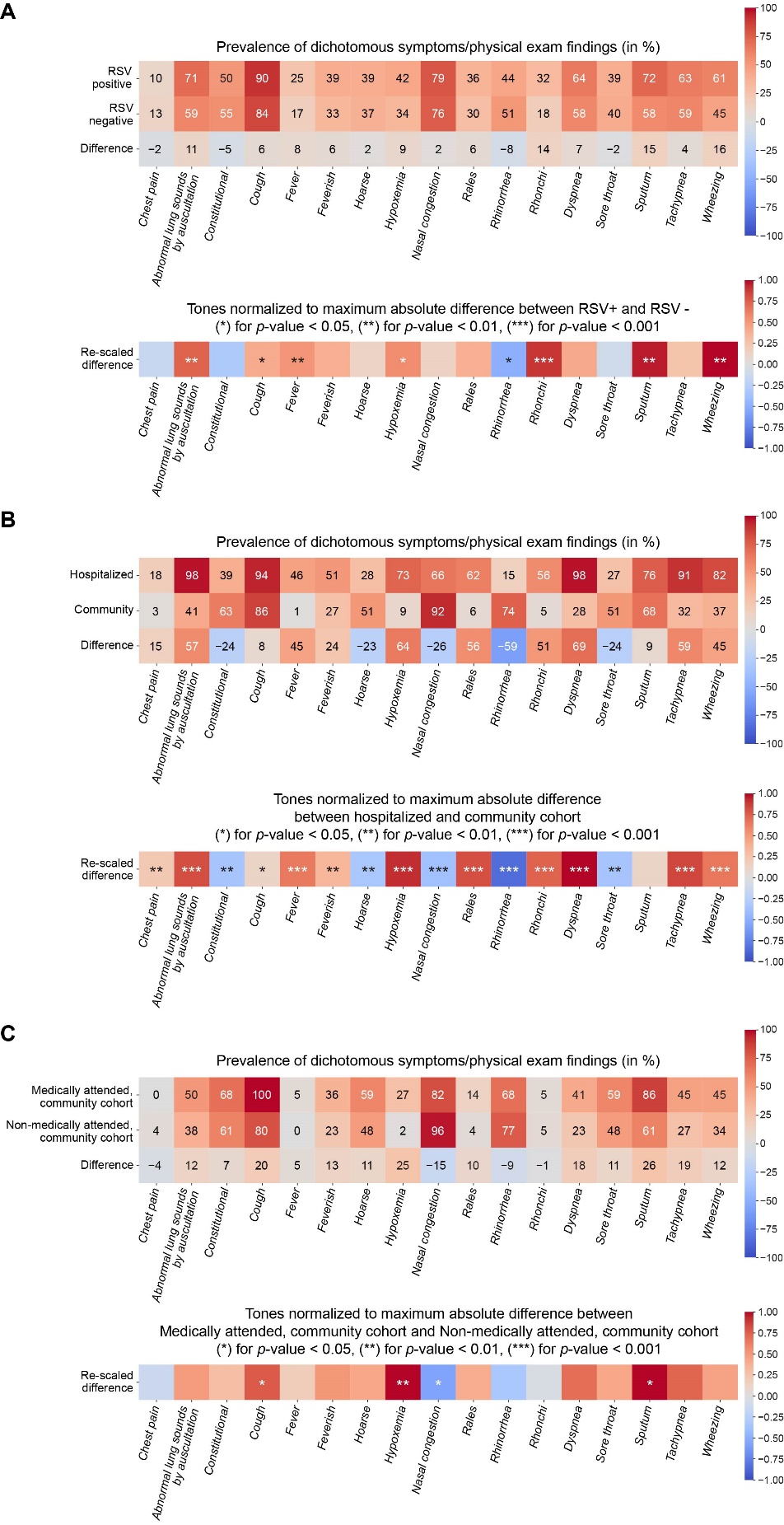

#### **Supplemental Figure 3.** Feature importance of machine learning model input variables to predict severe RSV-LRTD among RSV-positive patients (n=163)

#### Abbreviations: CHF, congestive heart failure; COPD, chronic obstructive pulmonary disease; LRTD, lower respiratory tract disease; RSV, respiratory syncytial virus.

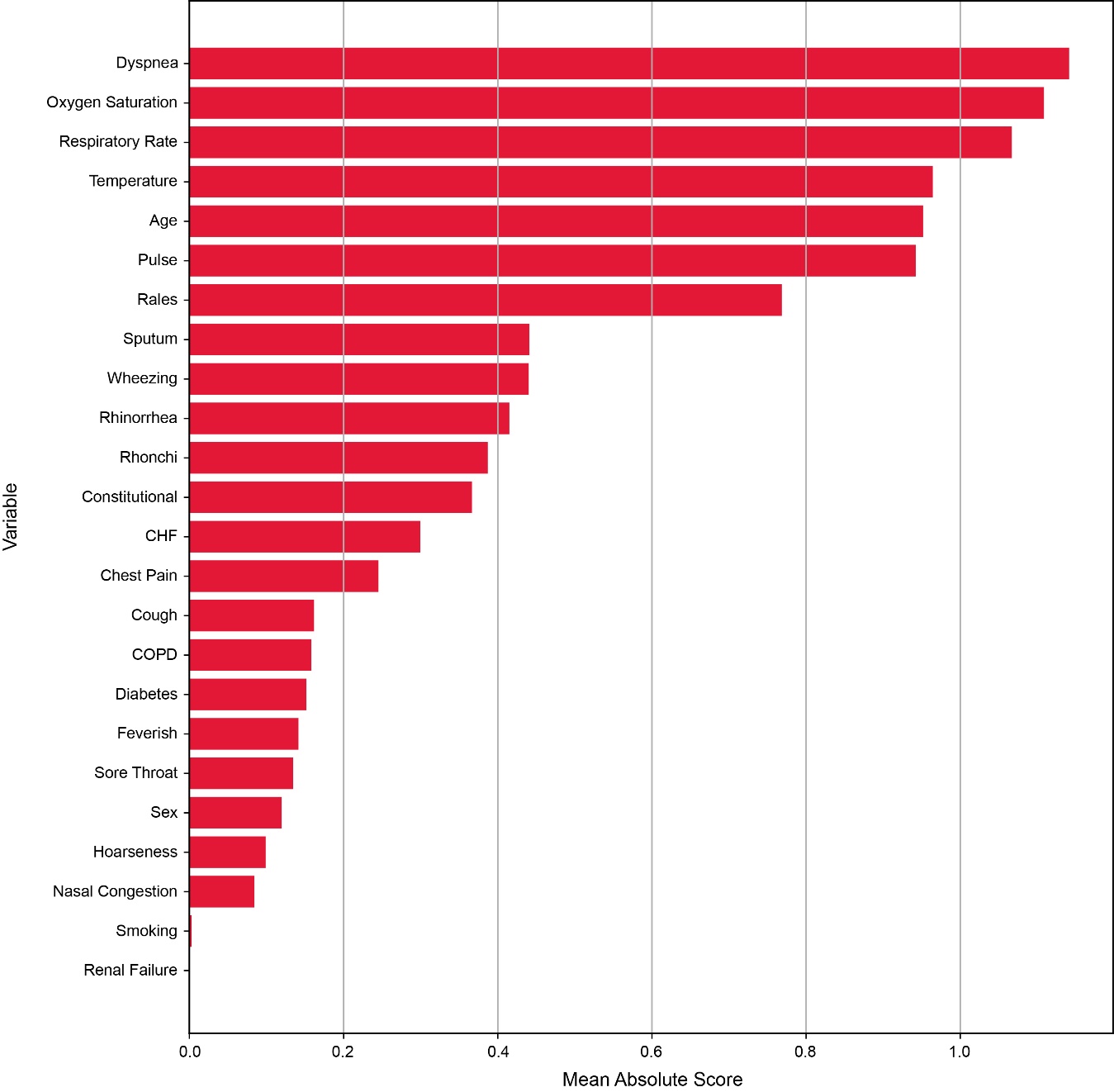

#### **Supplemental Figure 4.** Frequency of sign/symptom pairs. (A) Frequency of sign/symptom pairs in all participants who were RSV-positive (n = 163) or (B) RSV-negative (n = 836); (C) frequency of sign/symptom pairs in participants who were RSV-positive in the hospitalized cohort (n = 85) or (D) RSV- positive in the community cohort (n = 78); and (E) frequency of sign/symptom pairs in RSV-positive participants in the community cohort who were medically attended (n = 22) or (F) non–medically attended (n = 56).

The color legend represents empirical probability of joint appearance of sign/symptom pairs (in %). Color scale is between 0% (dark blue) and 100% (light yellow), with the “yellower” colors representing higher values.

Abbreviation: RSV, respiratory syncytial virus.

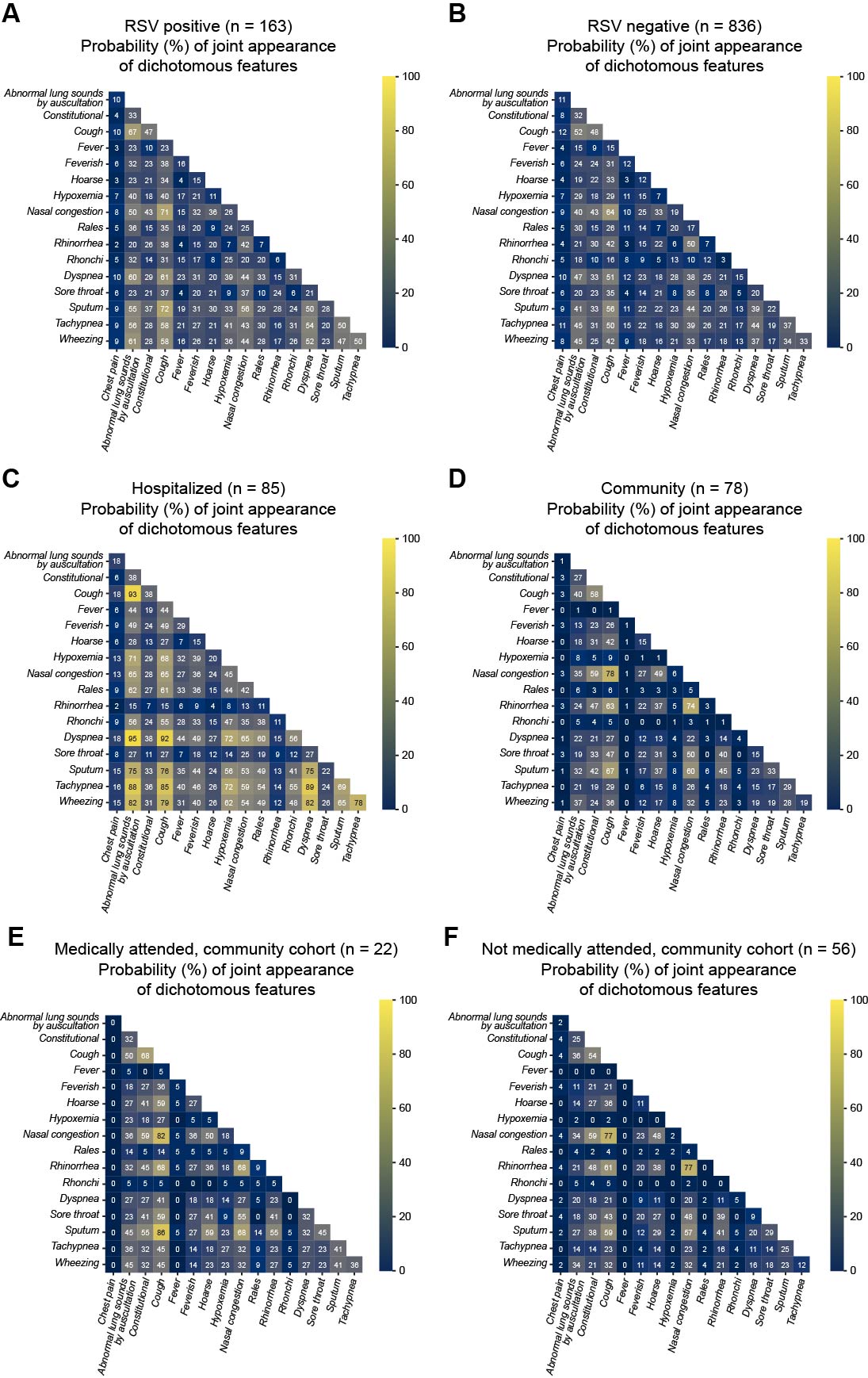

**Supplemental Figure 5.** Difference in the probability of sign/symptom pairs. (A) between participants who were RSV-positive and RSV-negative, (B) between the hospital and community cohorts, and (C) between medically and non-medically attended RSV-positive participants. The color legend represents difference of probability of joint appearance of sign/symptom pairs (in %). Color scale is between the lowest difference and the highest difference. Positive differences are shown in red and negative differences are shown in blue. The darker the red color, the higher the positive difference, while the darker the blue color, the lower the negative difference. Abbreviations: RSV, respiratory syncytial virus.

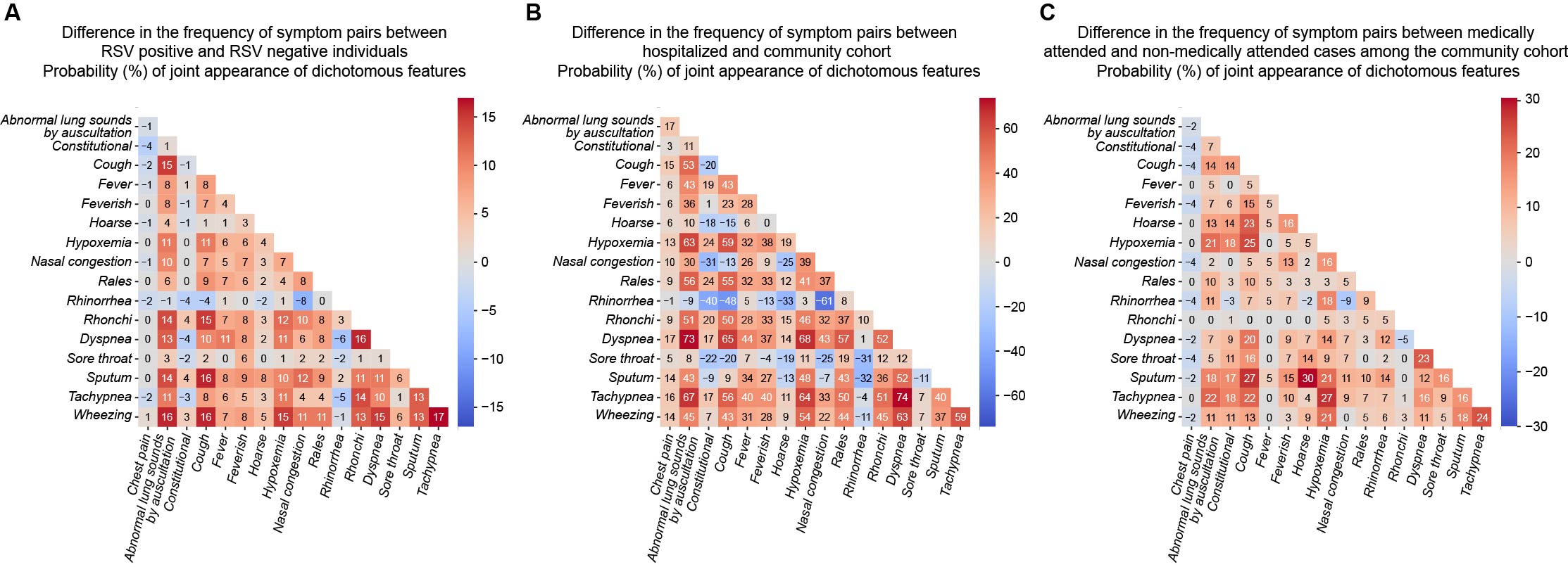
